# Location-, intensity-, and frequency-optimized epidural stimulation restores hand function after complete spinal cord injury

**DOI:** 10.64898/2026.04.07.26349471

**Authors:** Jeonghoon Oh, Alexander G. Steele, Michelle Scheffler, Catherine Martin, Jony Sheynin, Valerie A. Dietz, Alejandra Valdivia-Padilla, Jenny Dinh, Argyrios Stampas, Radha Korupolu, Christof Karmonik, Timea M. Hodics, Yevgeniy Freyvert, Michael Manzella, Amir H. Faraji, Philip J. Horner, Dimitry G. Sayenko

## Abstract

Cervical spinal cord injury (SCI) causes profound and persistent loss of hand function, and effective neuromodulation strategies remain limited. We report the first-in-human implantation of a 32-contact cervical epidural paddle array in two individuals with severe chronic SCI. Individualized motor pool recruitment maps, derived from systematic bipolar and multipolar configurations, enabled person-specific stimulation parameters. Optimized stimulation restored volitional hand opening, closing and coordinated upper-limb movements that were previously unattainable. This approach achieved a >91% success rate in complex reach-grasp-lift-release sequences, supported by substantial gains in range of motion, grip, and pinch strength. Electrophysiological and kinematic analyses demonstrated parameter-dependent, selective recruitment of flexor and extensor motor pools. Personalized stimulation programs integrated with goal-directed activities enabled functional hand use in home and community settings, sustained over several months of continued autonomous use. These findings establish a mechanistically grounded and translational framework for restoring upper-limb function after chronic severe SCI.

## Introduction

Spinal cord injury (SCI) imposes profound and lifelong physical, emotional, and societal burdens on affected individuals and their families. Cervical injuries account for nearly 60% of all cases^1^, and frequently result in tetraplegia, wherein loss of independence in essential daily activities such as eating, dressing, and personal hygiene is a common factor. Individuals with tetraplegia consistently rank restoration of arm and hand function as their highest rehabilitation priority^2,3^, as even modest improvements can substantially enhance autonomy and quality of life. Despite advances in rehabilitation and assistive technologies, meaningful recovery remains limited, particularly in individuals with chronic motor and sensory complete injuries.

Neuromodulation of spinal circuits via electrical stimulation has emerged as a promising strategy to restore motor function after SCI^4–7^. Lumbar epidural spinal stimulation (ESS) has enabled voluntary movement, standing, and stepping in individuals with paraplegia^8–10^, likely through the convergence of descending drive and afferent feedback onto spinal interneuronal networks that engage locomotor-related spinal circuits. However, upper-limb control differs fundamentally from locomotion. Arm and hand movements require selective activation of cervical motor pools and depend strongly on corticospinal drive, with less reliance on intrinsic rhythmic spinal mechanisms ^11^. As a result, neuromodulation strategies that prove effective in the lumbar cord do not directly translate to the cervical cord without spatial and temporal precision in circuit engagement. This distinction has contributed to the long-standing assumption that individuals with motor and sensory complete cervical injuries may lack sufficient descending control to benefit from stimulation, often leading to their exclusion from restorative neuromodulation approaches targeting upper-limb function.

Cervical transcutaneous spinal stimulation (TSS) has facilitated encouraging improvements in individuals with incomplete injuries^12–14^, but current TSS approaches lack the spatial resolution required to selectively engage specific motor pools^15,16^, and provide limited mechanistic insights. Moreover, previous studies with TSS have largely excluded individuals with motor and sensory complete injuries or reported highly variable outcomes^13,17,18^, leaving unresolved whether spinal circuits below a clinically complete lesion can be functionally leveraged. Cervical ESS offers the potential for location-specific engagement of spinal motor circuits and for modulating interactions between residual descending inputs and segmental circuitry to support volitional control^11^. An early feasibility study demonstrated acute re-engagement of upper-limb movements after chronic motor complete SCI^17^, while recent work in stroke showed that segment-specific cervical stimulation can enhance upper-limb muscle strength and dexterous hand function^19^. However, the physiological mechanisms governing these effects and the stimulation strategies required to restore function in complete cervical injuries remain poorly defined, particularly given substantial inter-individual variability in injury, anatomy, and residual connectivity.

Here, we used a surgically implanted 32-channel paddle array placed over the cervical enlargement to generate individualized motor pool maps and identify stimulation configurations that selectively engage upper-limb motoneurons. In two individuals with motor and sensory complete SCI, targeted stimulation produced salient and reproducible improvements in volitional hand opening and closing, precision pinch, manipulation of small and heavy objects, and trunk stability, substantially exceeding baseline capabilities. Continued in-home use of individualized stimulation paradigms paired with goal-directed activities was associated with progressive gains in relevant daily activities. Electrophysiological and kinematic analyses revealed that stimulation intensity, location, and frequency systematically shape spinal circuit excitability and modulate interactions with descending pathways. In particular, frequency-dependent synaptic mechanisms and intensity-dependent recruitment thresholds enabled functional separation of overlapping motor pools, including a bias in wrist flexor versus extensor recruitment that supported person-specific stimulation strategies. Together, these findings define foundational mechanistic principles for restoring complex upper-limb function through personalized cervical spinal stimulation.

## Results

### Participant characteristics and study design

We report the first two participants enrolled in an ongoing clinical study evaluating the effectiveness of cervical ESS in upper-limb functional restoration after chronic SCI. The study followed a staged, within-subject design that transitioned from baseline clinical characterization to exploratory mapping, followed by functional optimization, and ultimately home use (**Fig. 1a**).

**Figure 1.**
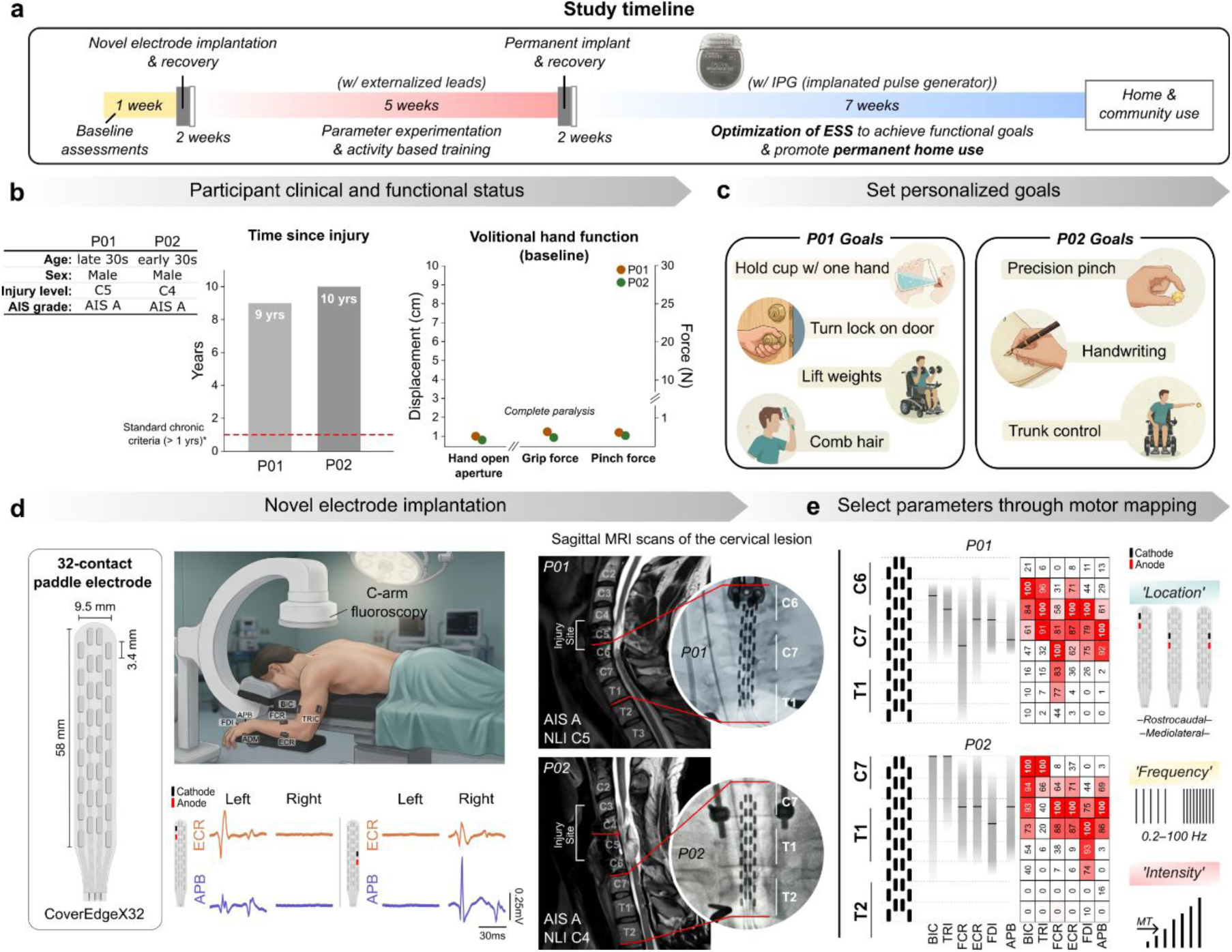
Experimental framework. **a**. Timeline of the staged, within-subject study design, progressing from baseline assessments and electrode implantation with externalized leads to permanent implantable pulse generator (IPG) placement and independent home use. **b.** Participant clinical and functional status. Both participants (P01, P02) presented with chronic^20^, motor and sensory complete cervical spinal cord injury (AIS A) and exhibited near-zero baseline grip and pinch force and displacement during attempted hand movements. **c.** Personalized functional goals were established prior to the intervention to guide parameter optimization. P01 focused on power grasp and complex hand functions, while P02 prioritized precision pinch and trunk stability. **d.** Surgical implantation of a 32-contact epidural paddle electrode array. Intraoperative C-arm fluoroscopy and sagittal magnetic resonance imaging (MRI) confirm array positioning over the cervical enlargement, caudal to the lesion site (C6–T1 for P01; C7–T2 for P02). Representative intraoperative electromyography (EMG) traces demonstrate lateralized upper-limb muscle activation. **e.** Individualized motor pool mapping. Heatmaps display the spatial recruitment profiles of upper arm, forearm, and intrinsic hand muscles across the electrode array. Stimulation location, frequency, and intensity were systematically tuned to achieve selective motor pool engagement.

Both individuals had traumatic, motor and sensory complete cervical injuries sustained approximately a decade prior to enrollment, with clinical presentations consistent with profound loss of voluntary hand function (**Fig. 1b**). Despite preserved proximal activity in muscles such as biceps brachii, the distal motor control (including wrist and intrinsic hand muscles) was largely absent, resulting in severe impairment of grasp, pinch, and complex hand functions. To ensure translation of the study interventions into person-centered outcomes, participants identified individual goals as a critical component of the trial. These goals guided the systematic optimization of stimulation (**Fig. 1c**).

After the baseline assessment, a 32-contact epidural paddle array was implanted over the cervical enlargement, near the caudal border of the lesion (C6–T1 in P01; C7–T2 in P02) to maximize access to upper-limb motor pools (**Fig. 1d**). Intraoperative mapping revealed consistent mediolateral selectivity, with preferential ipsilateral activation, and a rostrocaudal recruitment gradient. Following post-operative recovery, participants underwent repeated electrophysiological assessments to characterize stimulation-evoked motor responses across the electrode array (**Fig. 1e**). However, spatial targeting alone proved insufficient in resolving the dense overlap of forearm and hand motoneurons. Instead, selective control emerged from the interaction of electrode configuration with stimulation frequency and intensity. This parametric integration enabled the unique functional separation of overlapping motor pools and established the foundation for personalized, goal-directed stimulation paradigms (**Supplementary Figure 1**).

### Selective modulation of upper-limb motor pools to restore hand function

Following a two-week postoperative recovery, we iteratively optimized stimulation parameters using a structured decision framework (**Supplementary Figure 1**). This process revealed that motor pool recruitment was highly dynamic, shaped by the three-dimensional interplay of stimulation location, frequency, and intensity. Initial screening of cathode-anode pairs identified configurations that preferentially engaged task-relevant muscles. The subsequent refinement (often requiring multipolar arrangements) further enabled critical precise control of agonist-antagonist balance. Ultimately, functional selectivity emerged from the interaction of spatial targeting with frequency- and intensity-dependent modulation, rather than electrode configuration alone. This integrated, parameter-specific approach used throughout the intervention for selected functional activities was defined as the *Optimized ESS* configuration.

The systematic investigation of this physiological selectivity in both participants translated into meaningful motor recovery and profound functional gains. Of critical importance, Optimized ESS consistently enabled hand opening and closing, which were entirely absent without stimulation (**Fig. 2a-c**). These movements were effective, consistent, and repeatable, as well as highly parameter-dependent, indicating that functional output reflected selective engagement of underlying motor pools rather than nonspecific excitation. Differences between array positioning further highlighted the importance of anatomical targeting. In P01, partial coverage of C6 segments, corresponding to forearm extensor motor pools, was associated with robust return of volitional hand opening, whereas this effect was less pronounced in P02 (**Fig. 2d**). Electromyographic (EMG) recordings revealed marked amplification and coordination of forearm and intrinsic hand muscle activity during stimulation, with clear differentiation between hand opening and closing phases (**Fig. 2e**). At the behavioral level, stimulation transformed fragmented or ineffective attempts into coordinated movements, as reflected by expanded hand aperture (**Fig. 2b**), increased active range of motion, and substantially higher peak velocities in both participants (**Fig. 2c**). Beyond ESS-induced movements (**Fig. 2a**), Optimized ESS further enabled the restoration of fluid, volitional motor control. Notably, a constant “hand-open” configuration enabled full volitional hand closing-opening cycles (**Fig. 2f**). These cycles included active hand closing against a prevailing extensor-biased stimulation, followed by reopening. Although closing movements under hand-opening configurations were slower than those observed with hand-closing configurations, their consistent and reproducible execution indicates that descending volitional commands can effectively overcome stimulation-imposed activation patterns and reconfigure the net motor output. These observations provide direct evidence of restored supraspinal control in the presence of Optimized ESS. By contrast, the hand-closing configuration did not efficiently produce hand opening. Together, these findings demonstrate that Optimized ESS enables selective engagement of overlapping motor pools to generate task-specific, volitional hand movements, with functional outcomes determined by the interaction between stimulation parameters and paddle array positioning.

**Figure 2.**
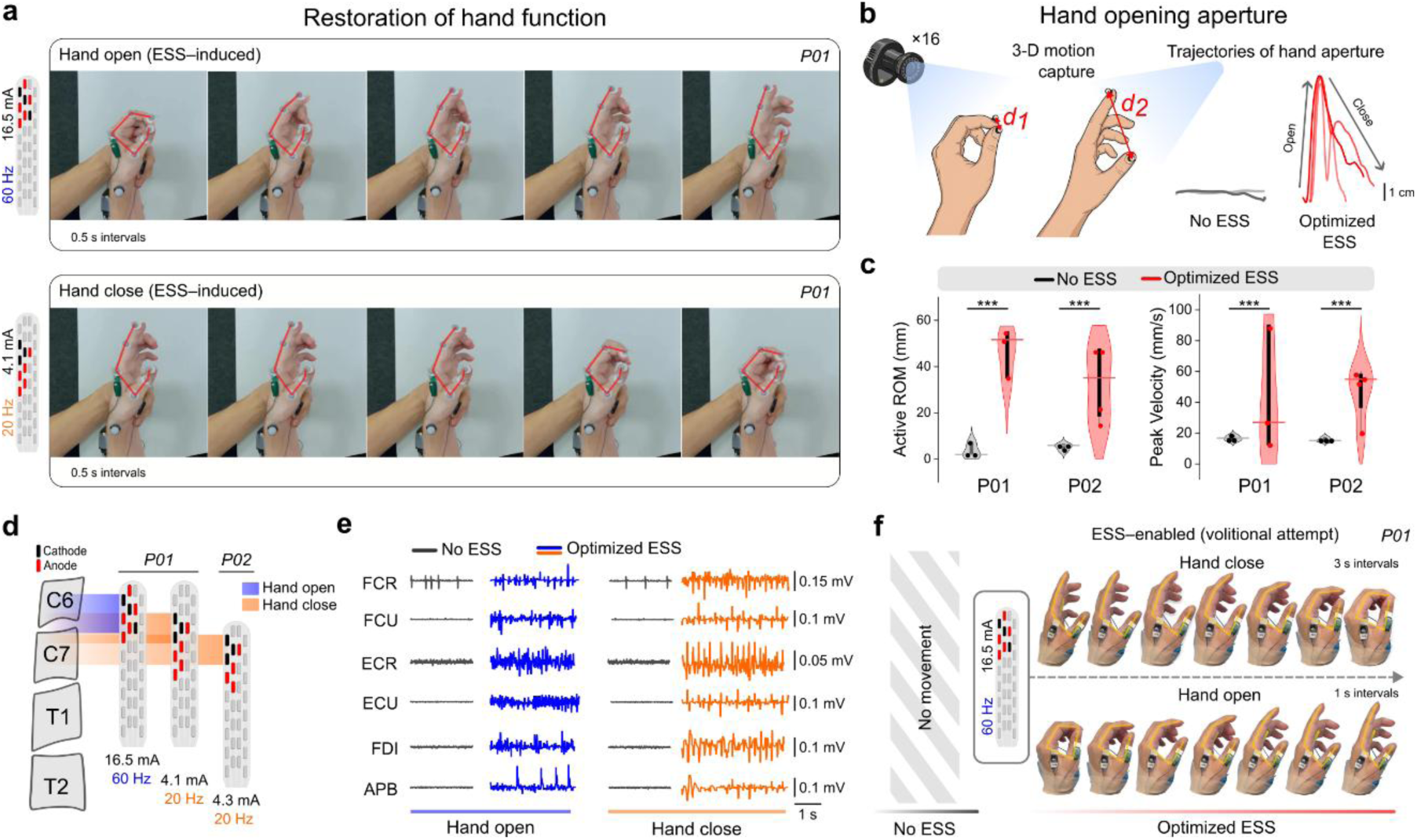
Restoration of hand function. **a.** Sequential snapshots (0.5-s intervals) of P01 performing volitional hand opening and closing during ESS-induced facilitation. Parameter-specific configurations were used for finger extension (rostral stimulation location, higher intensity, 60Hz) and flexion (caudal stimulation location, lower intensity, 20Hz). Red lines visualize hand opening aperture. **b.** Schematic of 3D motion capture setup using 16 infrared cameras to measure hand aperture, defined as the Euclidean distance between thumb and index fingertips. Resulting trajectories contrast No ESS (black) and Optimized ESS (red) conditions during a hand-opening cycle. **c.** Quantitative kinematic performance for P01 and P02. Violin plots with individual data points (circles) contrast No ESS (grey) and Optimized ESS (red) conditions for active range of motion (ROM) (left) and peak velocity (right) of the hand open-close cycle. Circles represent individual trials. Significant differences are indicated by asterisks (***P < 0.001; statistical tests are defined in the Methods). **d.** Projection of the 32-contact paddle arrays onto the cervical vertebrae for P01 and P02, illustrating the individualized cathode (black) and anode (red) configurations optimized for hand opening (blue shading, C6 level) and closing (orange shading, C7 level). **e.** Representative raw EMG traces of forearm and intrinsic hand muscles and hand aperture trajectories during attempted hand movements. Forearm and intrinsic hand muscles exhibit robust, parameter-specific activation only under Optimized ESS (colored traces, blue for opening, orange for closing), while No ESS (black) traces are silent. **f.** Restoration of volitional motor control during ESS-enabled hand close-open cycle. Sequential snapshots (intervals as indicated) demonstrate P01 performing active, volitional hand closing and opening cycles enabled by a constant, hand-opening configuration. No volitional movement was observed without stimulation (No ESS).

### Distinguishing specific from non-specific ESS effects

Systematic exploration of the implanted 32-contact array revealed that cervical ESS can generate a vast repertoire of upper-limb muscle activation patterns, many of which do not translate into meaningful movement. To resolve this ambiguity, we coupled EMG with direct quantification of joint-level output using a custom device measuring isometric forces at the wrist (**Fig. 3a**). This confirmed our earlier observations that overlapping motor pools frequently produced co-activation of antagonists, overpowering functional movement attempts despite robust muscle activity. However, we uncovered a unifying organizing principle: frequency-dependent modulation of motor pool recruitment. With electrode configuration held constant in the resting state, stimulation frequency alone reshaped recruitment dynamics and defined the direction of net force output (**Fig. 3b**). Lower frequencies (10–30 Hz) preferentially engaged flexor motor pools, including the flexor carpi radialis (FCR) and flexor carpi ulnaris (FCU), whereas higher frequencies (40–70 Hz) recruited extensors, such as the extensor carpi radialis (ECR) and extensor carpi ulnaris (ECU). This reciprocal tuning was consistent across participants (all P < 0.001), establishing frequency as a control parameter that facilitates agonist-antagonist balance and defines the direction of net force output.

**Figure 3.**
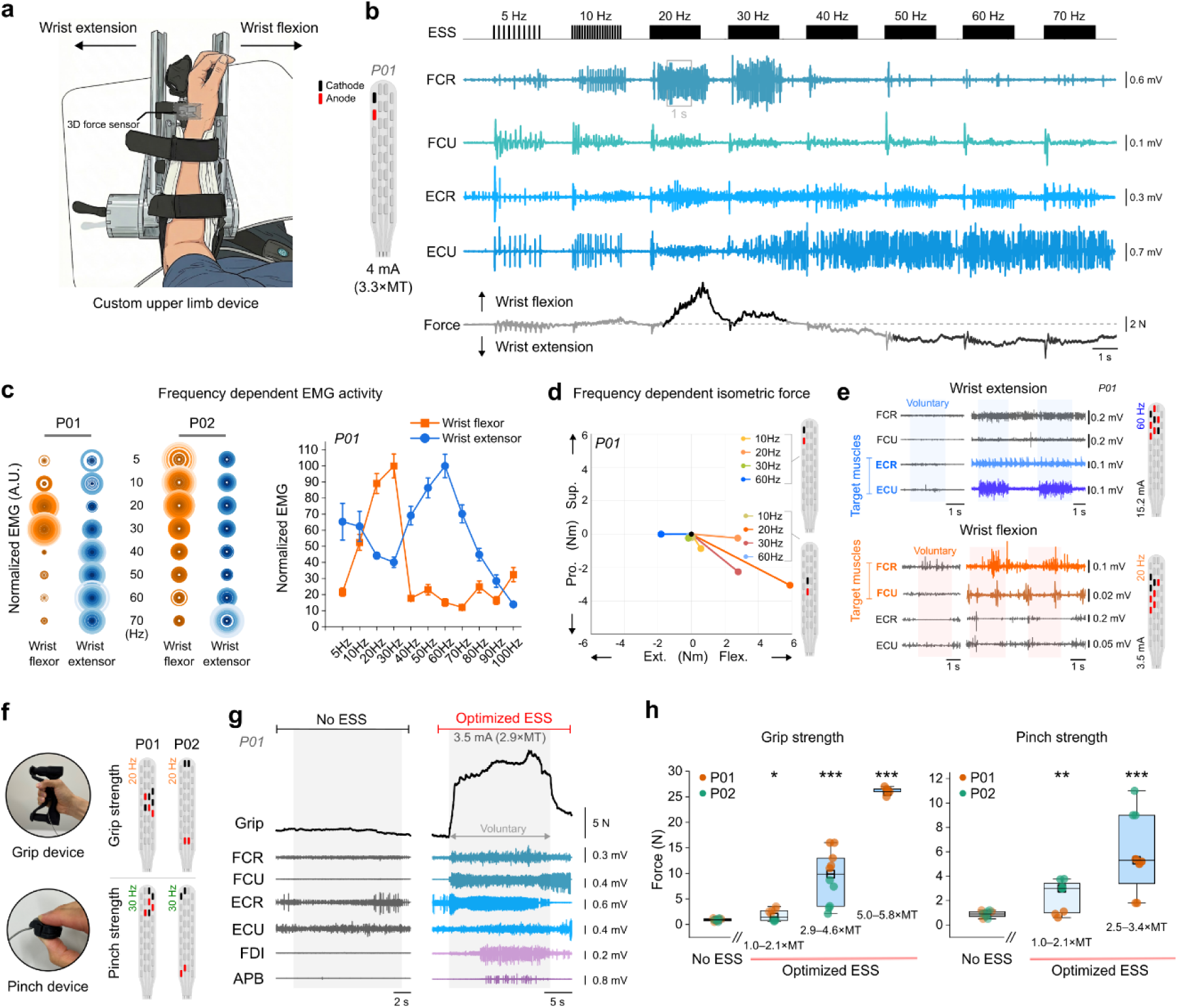
Frequency- and intensity-dependent modulation of upper-limb movement. **a.** Schematic of the custom isometric device used to quantify wrist flexion and extension forces. **b.** Representative electromyography (EMG) and force traces from P01 during a frequency sweep (5–70 Hz) at a constant stimulation amplitude (4 mA). Lower frequencies (e.g., 20 Hz) preferentially recruited wrist flexors (FCR, FCU) and generated flexion force, whereas higher frequencies (e.g., 60–70 Hz) shifted recruitment to wrist extensors (ECR, ECU) and produced extension force. **c.** Quantification of frequency-dependent motor pool recruitment. Left: Bubble plots representing the shift in magnitude of flexor (orange) and extensor (blue) EMG activation for P01 and P02. Right: The line-scatter plot shows normalized EMG activity for P01 across a frequency range of 5–100 Hz. Flexor activation (orange) peaks at 20–30 Hz, while extensor activation (blue) exhibits a distinct peak at 60 Hz, demonstrating the frequency-dependent selectivity used to optimize motor pool recruitment. **d.** 2D force vector map illustrating shifts in isometric wrist torque directionality (flexion/extension and pronation/supination) modulated by stimulation frequency and electrode location in P01. **e.** Representative EMG traces contrasting voluntary effort with Optimized ESS during wrist extension (top) and flexion (bottom). Optimized ESS provided robust facilitation of target muscle activation, whereas EMG activity remained largely absent during voluntary attempts without stimulation (No ESS). **f.** Custom devices for measuring grip and pinch strength, alongside participant-specific optimized electrode configurations (anode in red, cathode in black). **g.** Representative traces during an attempted voluntary grip task (P01). Optimized ESS enabled sustained voluntary force generation and robust, coordinated activation of forearm and intrinsic hand muscles compared to the No ESS condition. **h.** Box plots detailing maximal grip and pinch forces for P01 (orange) and P02 (green) under No ESS and varying intensities of Optimized ESS (expressed as multiples of motor threshold, ×MT). Boxes span the interquartile range, horizontal lines indicate medians, and whiskers represent data limits. Individual data points are overlaid. Asterisks denote statistical significance compared to No ESS (*P < 0.05, **P < 0.01, ***P < 0.001; see Methods for details of statistical analyses).

These frequency-dependent effects translated directly into specific movement patterns. During attempted wrist flexion, ESS at 20 Hz selectively amplified flexor activation, producing coordinated flexion forces that were absent without stimulation (**Fig. 3c**). By contrast, 60 Hz stimulation facilitated extension, driving phasic activation of extensor pools while limiting flexor co-activation. Notably, this bidirectional control was participant-specific: P01 exhibited clear flexion-extension patterns while P02 remained flexor-dominant, with suprathreshold stimulation preferentially recruiting flexor circuits and limiting extension. Across conditions, Optimized ESS significantly increased EMG in targeted flexor or extensor groups compared to No ESS, with corresponding shifts in force directionality (**Fig. 3d-e**).

Stimulation parameters were further individualized to maximize functional outputs, with location-specific configurations defining the spatial recruitment of motor pools. Optimized ESS enabled substantial gains in power grasp, increasing maximal force from near-zero levels (∼1.2 N) to functionally relevant magnitudes (up to 27.3 N in P01; P = 0.006), sufficient for object manipulation (**Fig. 3f-h**). Increasing stimulation intensity produced graded amplification of muscle activation and grip force generation, with reproducible gains across intensities reflecting coordinated recruitment of wrist and intrinsic hand muscles (**Fig. 3h**). Crucially, this robust functional enhancement required precise suprathreshold titration; stimulation at mere motor threshold (MT) or high-amplitude stimulation at off-target locations failed to elicit meaningful motor output (**Supplementary Figure 2**). In P02, early improvements were masked by elevated flexor tone, but longitudinal assessments revealed progressive increases in grip strength, underscoring the importance of joint stabilization for effective force transmission (**Fig. 3h**).

Optimized ESS also enabled precision control. Tripod pinch, absent under No ESS, emerged with Optimized ESS in both participants, with pinch forces reaching 5–9 N (up to a 9-fold increase) and accompanied by selective activation of intrinsic hand muscles including the first dorsal interosseous (FDI), abductor pollicis brevis (APB), and abductor digiti minimi (ADM) (all P < 0.001). These effects supported fine manipulation tasks, indicating that Optimized ESS organizes spinal network activity into coordinated, task-specific motor output.

### Optimizing ESS for complex functional upper-limb movements

Building on the frequency-, intensity-, and location-specific control of motor pools (**Fig. 3**), we tested whether these principles could be combined to support coordinated, multi-phase movements required for daily activities (**Fig. 4**). Without stimulation, both participants exhibited profound motor deficits. Volitional hand opening was entirely absent, and attempts to manipulate the can of soda relied on suboptimal compensatory strategies (e.g., tenodesis grasp) that failed to achieve functional digit integration or generate sufficient grasp force for lifting.

**Figure 4.**
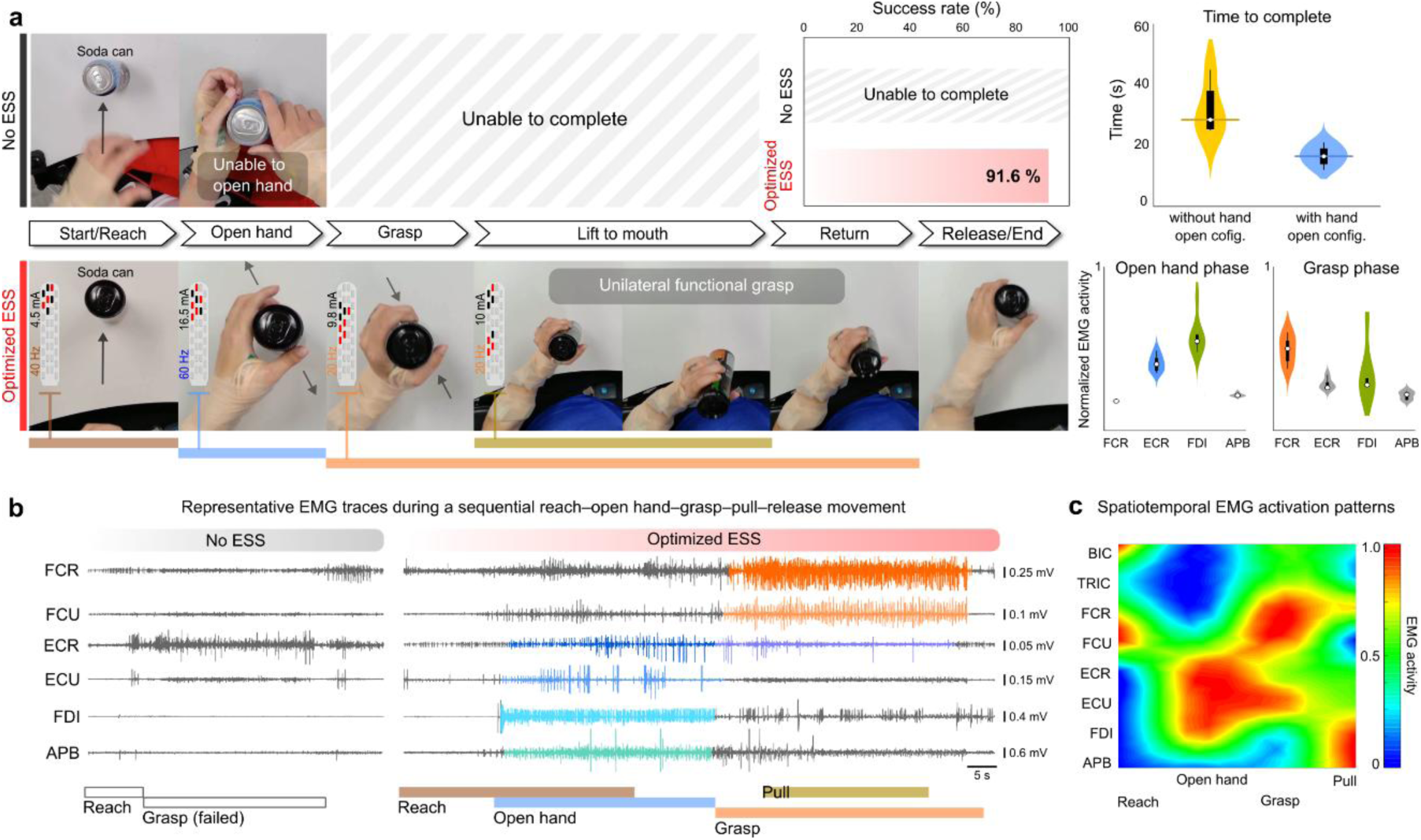
Optimized ESS restores complex upper-limb movements. **a.** Sequential snapshots comparing performance of a functional soda can drinking task. Without stimulation (No ESS, top), the participant fails the task due to an inability to volitionally open the hand and insufficient grasp strength to lift the object. Optimized ESS (bottom) enables successful completion of the multi-phase task (reach, open hand, grasp, lift to mouth, return, and release) via dynamic switching of phase-specific stimulation parameters (i.e., location, frequency, and intensity of stimulation). Top right: Without stimulation (No ESS), the task was impossible to complete, whereas Optimized ESS enabled a 91.6% success rate. Violin plots demonstrate that incorporating a dedicated ‘hand-open’ stimulation configuration reduced the time to complete the task compared to a ‘grasp’ configuration. Adjacent violin plots quantify normalized electromyography (EMG) activity across select forearm and intrinsic muscles during the ‘open hand’ and ‘grasp’ phases. **b.** Representative raw EMG traces. The No ESS condition exhibits minimal to absent EMG activity, resulting in a failed grasp. By contrast, Optimized ESS drives robust, phase-locked muscle activation corresponding to the reach, open hand, and grasp/pull phases. **c.** Spatiotemporal heatmap of normalized EMG activity across proximal (BIC, TRIC), distal forearm, and intrinsic hand muscles during Optimized ESS, illustrating the coordinated, sequential muscle synergies required for successful object manipulation.

Optimized ESS by contrast facilitated the immediate restoration of hand functions that then enabled goal-directed movement. With Optimized ESS, P01 successfully completed a continuous reach-grasp-lift-return-release task, which he had not been able to perform since his injury (**Fig. 4a**). Successful completion relied on the dynamic, phase-locked switching of stimulation parameters to meet the opposing biomechanical demands of the task. Distinct configurations were sequentially triggered to support hand opening, grasp formation, sustained force generation, and object release.

When paired with volitional effort, this temporally targeted modulation markedly improved performance. Optimized ESS restored functional capacity, achieving a 91.6% success rate. In particular, incorporating a dedicated ‘hand-opening’ component within the Optimized ESS parameters significantly reduced total task completion time when compared to a paradigm that only promoted grip (**Fig. 4a**). These behavioral gains were accompanied by phase-specific muscle recruitment, with coordinated activation of forearm and intrinsic hand muscles during the opening and grasp phases (**Fig. 4b-c**), demonstrating that Optimized ESS can organize sequential motor output to enable functional behavior.

### Translation to clinically meaningful upper-limb function

Optimized ESS translated directly into clinically measurable recovery of upper-limb function. To quantify these effects, we used standardized rehabilitation measures, specifically the Graded Redefined Assessment of Strength, Sensibility and Prehension (GRASSP version 2) and the International Standards for Neurological Classification of Spinal Cord Injury (ISNCSCI), at baseline and study completion. These were tested under No ESS and Optimized ESS conditions to dissociate stimulation-dependent effects from persistent effects (**Fig. 5**).

**Figure 5.**
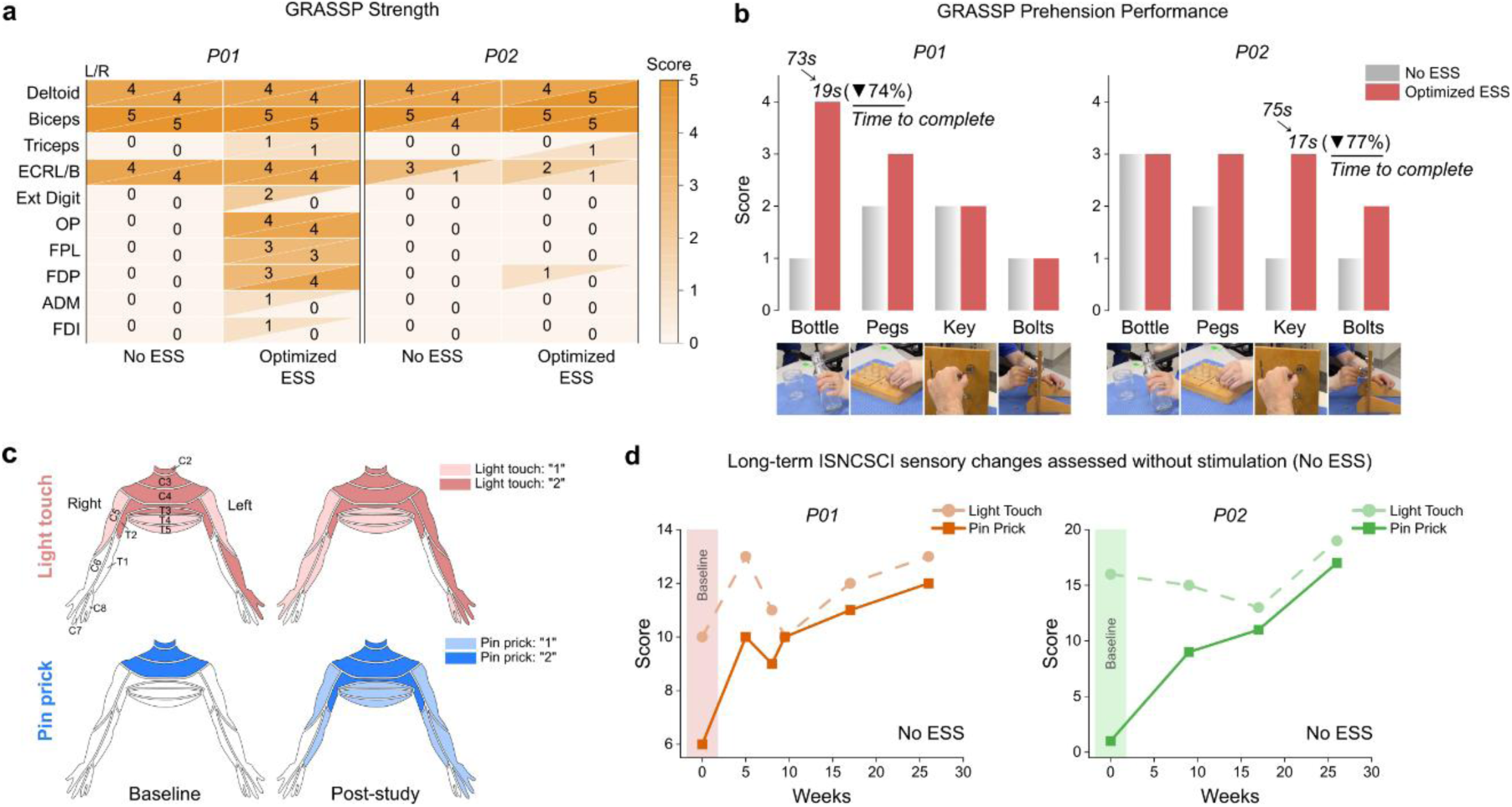
Optimized ESS improves standardized rehabilitation outcomes. **a.** Heatmap detailing bilateral (L/R) Graded Redefined Assessment of Strength, Sensibility and Prehension (GRASSP) strength subtest scores for P01 and P02. Optimized ESS facilitated immediate improvements, enabling the emergence of previously absent intrinsic hand and distal arm strength (scores > 0). **b.** GRASSP prehension performance. Bar graphs contrast No ESS (blue) and Optimized ESS (red) condition scores. Optimized ESS not only increased scores but also facilitated efficiency, highlighted by substantial reductions in task completion times (e.g., a 74% reduction during the ‘Bottle’ task for P01, and a 77% reduction during the ‘Key’ task for P02). **c.** Representative human dermatome maps (P01) illustrating the spatial expansion of ISNCSCI sensory scores. Shading intensity reflects the degree of preservation for light touch (red) and pinprick (blue) modalities (1 = altered, 2 = normal). **d.** Longitudinal evolution of ISNCSCI sensory scores (C5–T5 dermatomes). Both participants exhibited progressive, continuous increases in light touch (dashed lines) and pinprick (solid lines).

Optimized ESS produced immediate, clinically, and functionally meaningful gains. In P01, ISNCSCI upper-limb motor scores increased by 6 points, including the emergence of previously absent intrinsic hand muscle activity. P02 demonstrated a 2-point improvement in upper-limb motor scores. These changes were accompanied by robust increases in GRASSP strength and prehension, reflecting coordinated recruitment across proximal, distal, and intrinsic motor pools ( **Fig. 5a-b**).

Functionally, Optimized ESS enabled a transition from suboptimal compensatory strategies to successful, coordinated task execution incorporating more natural movement patterns. Additionally, Optimized ESS facilitated marked improvements in functional efficiency, yielding substantial reductions in task completion time (∼74–77%) during complex manual tasks (**Fig. 5b**).

Notably, these motor gains occurred in parallel with the expansion of light touch and pinprick sensation across upper-limb dermatomes. These sensory improvements progressed steadily over time and were documented even in the No ESS condition (**Fig. 5c-d**), substantiating the reorganization of distributed sensorimotor networks. Together, these findings demonstrate that Optimized ESS can rapidly restore clinically and functionally meaningful upper-limb function while unmasking latent sensorimotor capacity and early signs of circuit-level reorganization.

### Personalized paradigms restore meaningful daily activities

The functional gains achieved through Optimized ESS extended beyond laboratory performance, enabling restoration of meaningful, self-directed behaviors in home and community environments (**Fig. 6**). Following the study-based optimization, participants were provided with individualized stimulation programs for home and community use. This was achieved through use of a simplified interface, therefore enabling safe, efficient, and independent use during daily activities. With Optimized ESS, functions that were previously unattainable or severely impaired became feasible again. These include independent drinking, personal grooming, handwriting, precision manipulation (e.g., door-lock rotation), and upper-limb load-bearing tasks.

**Figure 6.**
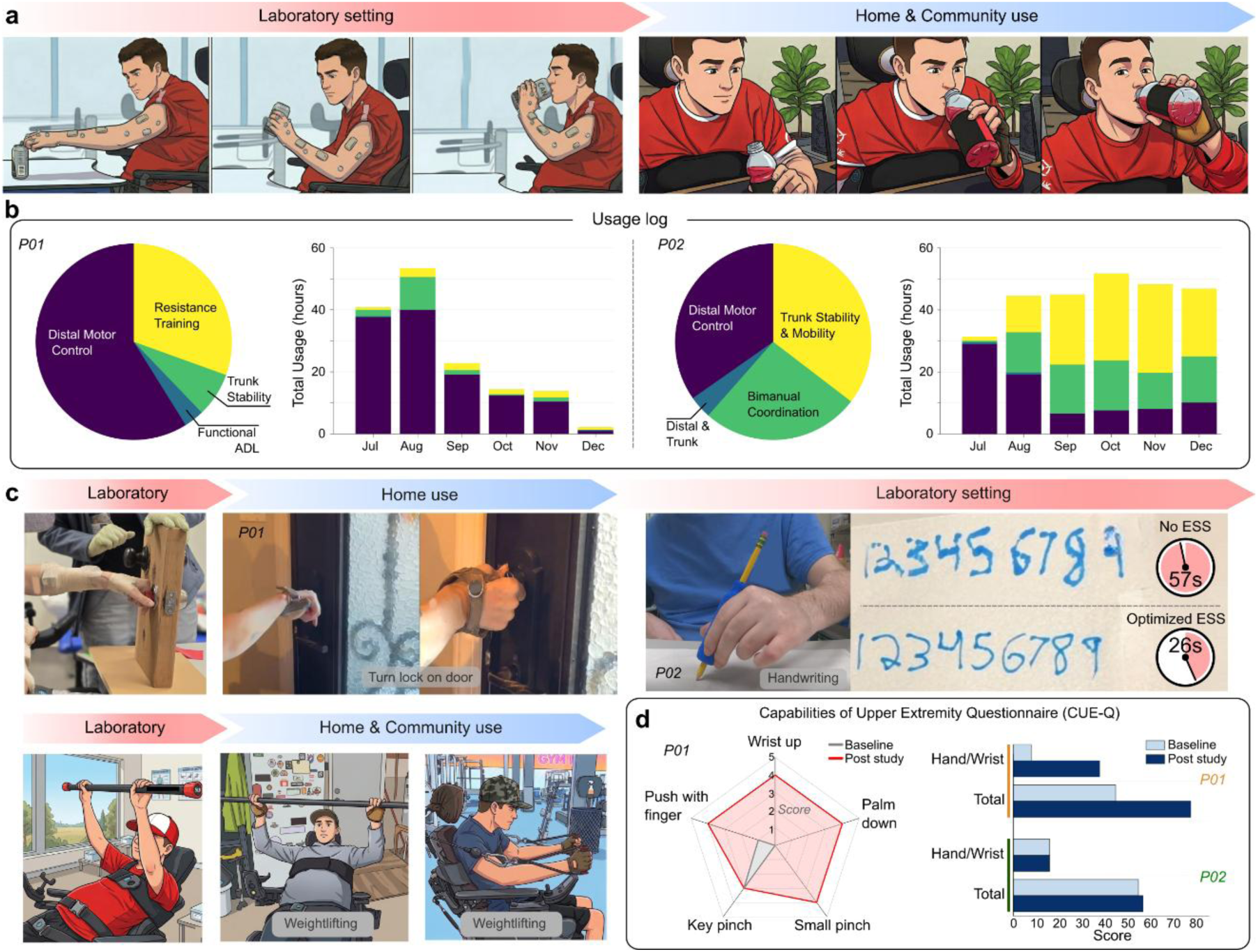
Personalized stimulation paradigms restore self-directed activities in home and community settings. **a.** Sequential illustrations depict P01 integrating laboratory practice into home and community environments. **b.** Longitudinal utilization. Usage logs for P01 (left) and P02 (right) detail ESS utilization across functional domains (pie charts) and total monthly hours of independent system use over the course of six months. **c.** Generalization of functional skills to home and community environments. Top left: ESS-enabled precision pinch during a door lock rotation task. Top right: Improved handwriting efficiency for P02, with a 54% reduction in execution time. Bottom left: Illustrations depict P01 performing a guided bench press in the laboratory, home, and a community gym, demonstrating progression of strength training across diverse settings. **d.** Standardized assessment of functional recovery. Left: Radar chart for P01 (Capabilities of Upper Extremity Questionnaire, CUE-Q) showing substantial gains across multiple hand/wrist domains from baseline (grey) to post-study (red). Right: Comparative hand/wrist and total CUE-Q scores for P01 and P02, confirming robust improvements in functional capabilities from baseline (light blue) to post-study (dark blue). In accordance with medRxiv policy, all photographs of participants have been removed and replaced with non-identifiable illustrations. The original materials are available from the corresponding author upon request, subject to institutional and ethical approvals.

To achieve their personalized goals (**Fig. 1c**), P01 utilized Optimized ESS to engage in progressive upper-limb loading outside the laboratory, progressing from assisted upper-limb exercises in his home to independent dumbbell and strength training workouts in a community gym. In P02, Optimized ESS restored stable precision control, enabling sustained pinch during forearm rotation and transforming handwriting into an effective, efficient and fluid execution of the task (reducing completion time by 54%, from ∼57 s to 26 s). In parallel, stimulation enhanced proximal trunk stability and reduced compensatory strategies, supporting efficient, coordinated reach-to-grasp movements in natural settings (**Supplementary Figure 3**).

Daily usage logs over the 6-month follow-up demonstrated the sustained, real-world integration of ESS into daily routines (**Fig. 6b**). Longitudinal analysis revealed distinct usage patterns aligned with individual functional priorities. P01 predominantly engaged the system for resistance training and distal motor control, with peak usage exceeding 50 hours/month during the initial phase of independent home use. By contrast, P02 exhibited progressive adoption, with monthly usage rising from ∼30 hours to a sustained plateau of nearly 50 hours, distributed across a broader set of functions, including distal control, trunk stability, and bimanual coordination tasks. These objective behavioral gains were mirrored by substantial improvements in self-reported function (Capabilities of Upper Extremity Questionnaire, CUE-Q) across both specific hand/wrist domains and total scores (**Fig. 6d**). Furthermore, daily usage logs over the follow-up period confirmed the active and sustained home and community use, suggesting integration into daily routines. Collectively, these findings demonstrate that personalized cervical ESS effectively translates selective motor pool control into restored upper-limb and hand function, ultimately leading to increased functional independence after chronic severe SCI.

## Discussion

This study provides direct evidence that Optimized ESS can restore coordinated, task-specific upper-limb function in individuals with chronic motor and sensory complete SCI. By combining individualized motor pool mapping with systematic control of stimulation parameters, we show that sub-lesional cervical circuits retain sufficient functional organization to generate meaningful motor output when appropriately engaged. Importantly, these effects extended beyond clinically measurable improvements in laboratory settings to self-directed activities in home and community environments, with early evidence of stimulation-independent gains.

To achieve these outcomes, we first addressed anatomical motor pool overlap that has historically constrained clinical translation of cervical spinal neuromodulation. Although the rostrocaudal organization of cervical motoneurons is well established^11,21^, translation has been limited by overlapping motor pool segmental representations and inter-individual variability. Here, subject-specific ESS optimization enabled selective engagement of distinct motor pools, including agonist-antagonist balance, which proved essential for coordinated wrist and hand control. Leveraging the spatial resolution, as well as frequency and intensity of ESS, we achieved reproducible recruitment patterns supporting wrist stability and functional hand opening-closing behaviors. By contrast to transcutaneous approaches that often produce diffuse activation^15,16,22,23^, ESS enabled focal, robust, and functionally relevant selectivity. Differences in implantation level across participants further highlight the need for individualized mapping to accommodate anatomical variability and optimize distal motor pool recruitment.

A central finding of this study is that Optimized ESS enables programmable control of motor output through the interaction of stimulation location, intensity, and frequency. Stimulation location determined the spatial recruitment of motoneurons, while intensity governed the magnitude and reliability of circuitry engagement. Frequency acted as a dynamic control parameter, shaping the balance between flexor and extensor activation and enabling bidirectional modulation of joint torque. Together, these parameters transformed stimulation from a nonspecific facilitatory input into a controllable system capable of shaping coordinated muscle synergies. This parametric framework allowed selective amplification of task-relevant motor pools while minimizing unwanted co-activation, a prerequisite for restoring functional hand movements.

By structuring parametric control over time, ESS coordinated motor output to support multi-phase movements. Complex tasks such as reach-grasp-lift-release required dynamic switching of stimulation parameters, with distinct configurations supporting individual movement phases. Crucially, while recent epidural stimulation studies in non-human primates^24^ and humans^19^ successfully augmented proximal reaching and flexor-dominant grip, restoring active hand opening has remained a critical limitation. Because cervical stimulation inherently biases toward flexor recruitment, eliciting voluntary hand extension was previously difficult to achieve^11,24^. We overcame this physiological constraint by integrating a dedicated ‘hand-open’ configuration, which proved vital for avoiding compensatory tenodesis and significantly reducing task execution time. This phase-specific control enabled participants to transition from fragmented, compensatory strategies to structured, goal-directed behavior, with marked improvements in success rate and execution speed. These findings extend prior reports of gross force augmentation by demonstrating that Optimized ESS can restore coordinated, sequential, task-locked motor patterns underlying functional upper-limb use in individuals with SCI.

Restoration of these motor patterns translated into quantifiable improvements on standardized rehabilitation measures. Gains in ISNCSCI motor scores and GRASSP strength and prehension reflected enhanced recruitment across upper arm, forearm, and intrinsic hand muscle groups, accompanied by substantial improvements in dexterity and task efficiency. Notably, both participants exhibited an expansion of light touch and pinprick sensation over time, even in the absence of ESS. While these changes should be interpreted cautiously, they are consistent with the engagement of distributed sensorimotor networks and suggest that repeated exposure to cervical ESS may promote broader changes in neural organization beyond immediate motor facilitation.

A defining advance of this work is the demonstration that ESS-enabled gains generalize beyond the laboratory to real-world environments. Participants independently deployed personalized stimulation programs outside the laboratory, restoring meaningful, self-directed upper-limb activities, such as drinking, handwriting, precision manipulation, and load-bearing, that were previously unattainable. These gains were integrated in daily routines and accompanied by improvements in self-reported function and sustained use in home and community settings. Together, these findings show that targeted cervical ESS extends beyond task-specific facilitation to support functional independence.

Critically, sustained engagement observed in our cohort addresses a major translational hurdle in SCI neuromodulation: long-term device adherence^25^. Evaluations of neuroprosthetic and electrical stimulation systems consistently identify declining use and device abandonment as primary barriers to clinical success^26^, often occurring when fixed stimulation parameters fail to provide meaningful daily utility once initial gains plateau^27^. In contrast, participants in our study maintained continued use of cervical ESS for six months post-study completion. We attribute this sustained adherence to our personalized optimization paradigm in which stimulation parameters were continuously tailored to achieve user-defined functional goals rather than selected solely for standardized clinical metric. By ensuring immediate, practical utility in real-world settings, this approach supports the long-term engagement necessary for meaningful functional improvements.

Our findings support a model in which targeted cervical ESS transforms spinal circuitry into a programmable substrate for motor control. Consistent with prior work, stimulation of the dorsal cervical cord leads to engagement of premotoneuronal and interneuronal networks, rather than direct motoneuron activation^28,29^. Within this framework, our data demonstrate that motor output is defined by the three-dimensional interplay of stimulation location, intensity, and frequency. Stimulation location determines the segmental networks and motor pools engaged, in accordance with cervical myotomal organization^19,21,30^. Stimulation intensity governs the reliability and magnitude of circuitry recruitment, with currents at or above motor threshold required to modulate ongoing voluntary activity^31–34^. Frequency further shapes interneuronal dynamics by tuning the balance of excitation and inhibition within these networks^29,35,36,37^. These parameter-dependent effects likely reflect the engagement of convergent premotoneuronal pathways, including propriospinal and reticulospinal circuits that remain partially preserved after SCI and can amplify residual descending input^36,38–40^ (**Supplementary Figure 4**). In this context, ESS does not simply increase excitability, but reconfigures the functional state of spinal networks, enabling the emergence of coordinated, task-specific motor output.

These findings establish a mechanistically grounded and clinically actionable framework for precision neuromodulation of the cervical spinal cord. By optimizing stimulation parameters with individual functional goals, this approach overcomes key limitations of prior studies, including variability in outcomes and limited functional specificity. Future work should extend this framework to larger cohorts, develop biomarkers to identify functions most likely to recover, and assess the durability of both stimulation-dependent and independent effects. Integrating closed-loop control, cortical stimulation, and task-specific rehabilitation may further enhance functional gains and promote activity-dependent plasticity.

## Conclusion

Personalized cervical epidural stimulation restores coordinated upper-limb function in chronic motor and sensory complete SCI by enabling selective, task-specific engagement of spinal sensorimotor networks. This optimized approach unlocks latent motor capacity and supports recovery from controlled laboratory settings to real-world independence. These findings define a new paradigm for spinal neuromodulation and establish targeted cervical ESS as a promising strategy to restore hand function and improve quality of life after severe cervical SCI.

## Methods

### Trial and participant information

All experimental procedures were approved by the Houston Methodist Research Institute Institutional Review Board (IRB) under an Investigational Device Exemption (IDE; protocol G230309). The study is registered at ClinicalTrials.gov (NCT06225245). Two male participants were enrolled in the study: Participant 01 (P01, late 30s years) and Participant 02 (P02, early 30s years), both with chronic, traumatic, motor and sensory complete cervical SCI at levels C5 and C4, respectively (**Fig. 1b**). Injuries were sustained during sporting accidents approximately ten years prior to enrollment and were classified as American Spinal Injury Association Impairment Scale (AIS) A according to the International Standards for Neurological Classification of Spinal Cord Injury (ISNCSCI)^41^. Both participants exhibited tetraplegia and functional impairments consistent with their clinical diagnoses and ISNCSCI assessments. At the time of enrollment, P01 showed left-side strength dominance but right-hand preference for fine motor skills, while P02 was left-side dominant. Participants provided written informed consent and were compensated for time and travel.

### Study design

Each participant underwent a structured, within-subject protocol spanning 17 weeks (**Fig. 1a**). Following baseline clinical assessments (Week 1), a 32-contact epidural paddle array was surgically implanted over the dorsal surface of the cervical spinal cord (Week 2). After a brief post-operative recovery period (Weeks 3–4), mechanistic assessments were conducted at Weeks 5, 8, and 11. Assessments included (i) spinal electrophysiology to map muscle-specific responses across stimulation configurations, frequencies, and amplitudes, and (ii) volitional output testing to quantify stimulus-enhanced force generation in upper-limb muscle groups under isometric conditions. In Week 9, each participant underwent surgical implantation of an internal pulse generator (IPG), followed by recovery and parameter refinement (Weeks 12–13) to optimize spatial and temporal targeting of upper-limb motor pools. A final evaluation was completed in Week 17, and the study concluded with a comprehensive clinical and functional assessment. Rehabilitation outcomes and long-term functional effects of ESS are beyond the scope of this report and will be presented in a separate publication.

### Surgical procedures

A 32-contact CoverEdge X32 paddle array (Boston Scientific, USA) was implanted by a certified neurosurgeon into the dorsal epidural space spanning C6–T1 in P01 and C7–T2 in P02, just caudal to the lesion site (**Fig. 1d**). A minimally invasive interlaminar approach was used, involving partial removal of the spinous process and separation of the ligamentum flavum to access the epidural space while preserving surrounding bony and ligamentous structures. The electrode array was secured to extension leads, which were externalized percutaneously at the T8 level, inferior to the right scapula, for connection to an external stimulator. Electrode positioning was confirmed intraoperatively using C-arm fluoroscopy and stimulation-evoked motor responses (**Fig. 1d**). A constant-current stimulator (DS8R, Digitimer Ltd., UK) was used to deliver controlled stimuli to verify selective activation of upper-limb muscles^19,42,43^. Participants remained under overnight hospital observation to monitor safety and ensure adequate recovery. In the second surgery (Week 9), a WaveWriter Alpha implantable pulse generator (Boston Scientific, USA) was implanted subcutaneously in the right subcostal region and connected to the previously implanted paddle array. Post-operative care followed the same monitoring and recovery protocol as the initial procedure.

### Clinical and functional assessments

Standardized clinical assessments, including ISNCSCI and GRASSP were conducted by trained therapists at each assessment. To evaluate functional relevance, participants performed specific motor tasks (e.g., precision grip, handwriting, drinking from a cup, and load-bearing activities). Complex multi-phase tasks were analyzed frame-by-frame via video recordings to quantify task success rates and time-to-completion (efficiency) under No ESS and Optimized ESS conditions. Capabilities of Upper Extremity Questionnaire (CUE-Q) scores and device usage logs were collected to evaluate self-reported functional improvements and home and community integration.

### Kinematic analysis

Kinematic data were captured using a 16-camera high-resolution 3D motion capture system (Vicon Valkyrie, Oxford, UK) sampling at 100 Hz, with data acquisition and hardware synchronization managed through Vicon Nexus v2 software. Temporal alignment between kinematic trajectories, surface EMG (Delsys Inc., USA), and force plate data was ensured via a Vicon Lock+ unit. To optimize tracking precision across varying anatomical scales, we utilized a dual-marker configuration: 9 mm diameter reflective markers were placed on the digits and hand to minimize mass-loading and inter-marker interference during fine motor tasks, while 14 mm markers were secured to the trunk and proximal upper-limb segments to ensure robust reconstruction.

Raw trajectories were exported as C3D files and imported into Visual3D (C-Motion, Germantown, MD, USA) for biomechanical modeling and analysis. Marker trajectories were filtered using a fourth-order, zero-lag, low-pass Butterworth filter (6 Hz cutoff), with segmental coordinate systems and joint centers defined from static calibration trials. Hand function was quantified by computing the 3D Euclidean distance (Dhand) between the distal phalanges of the thumb and index finger, serving as the primary metric for hand aperture during functional tasks (**Fig. 2b**). Simultaneously, postural control and trunk stability (**Supplementary Figure 3**) were evaluated by tracking spinal markers from C7 to S1 during upright sitting and dynamic reaching. These kinematic data were integrated with synchronized force plate (AccuGait-Optimized [ACG-O], AMTI, USA) to calculate the maximum anterior-posterior (A-P) center of pressure (COP) displacement and total COP path length, providing a comprehensive measure of dynamic balance and trunk stability.

### Electrophysiological assessment

Electromyographic (EMG) recordings were acquired bilaterally using wireless surface electrodes (Delsys Inc., USA) placed over key upper-limb muscles, including the biceps brachii (BIC), triceps brachii (TRI), flexor carpi radialis (FCR), flexor carpi ulnaris (FCU), extensor carpi radialis (ECR), extensor carpi ulnaris (ECU), first dorsal interosseous (FDI), abductor pollicis brevis (APB), and abductor digiti minimi (ADM). Signals were amplified using a Trigno amplifier (Delsys Inc., UK) and digitized at 2,000 Hz using a PowerLab data acquisition system (ADInstruments, Australia).

Spinally evoked motor potentials (SEMPs) were elicited using cervical ESS delivered via a constant-current stimulator (DS8R, Digitimer Ltd., UK). Monophasic pulses (0.1–0.5 ms) were applied at stimulation intensities ranging from 0 to 20 mA and frequencies from 0.2 to 100 Hz. Assessments were conducted both at rest and during controlled wrist flexion-extension movements to evaluate spinal circuitry excitability, motor pool recruitment, and modulation capacity. We tested bipolar cathode-anode pairs across the array to characterize both rostrocaudal (segmental) and mediolateral (lateralized) recruitment fields. For each muscle, response magnitude was plotted as a function of stimulation intensity, providing identification of activation thresholds and mapping of segmental motoneuron recruitment. These individualized maps guided the identification of optimal electrode configurations for targeted motor pool engagement (**Fig. 1e**). For testing of transsynaptic recruitment, two pulses were delivered at 0.2 Hz with a 30 ms inter-pulse interval.

Targeted ESS configurations were specifically mapped to motor pools to maximize activation of task-relevant muscles (e.g., flexors during wrist flexion), eliciting graded recruitment with clear thresholds and saturation at higher intensities. In contrast, the off-target ESS (sham) condition utilized caudal contacts (T1–T2) positioned outside functionally relevant segments. This configuration produced minimal or inconsistent upper-limb activation, serving as a control to dissociate non-specific stimulation effects (**Supplementary Figure 2**).

To assess how stimulation frequency modulates upper-limb motor pool recruitment, we performed a dedicated frequency-sweep protocol following completion of electrode-site mapping. For each participant, a task-relevant electrode configuration (identified during rostrocaudal/mediolateral mapping) was selected and held constant while amplitude and frequency were systematically varied. Stimulation consisted of trains of monophasic pulses (0.2 ms) delivered at supra-threshold intensities sufficient to evoke consistent SEMPs. Frequencies were tested sequentially at 5, 10, 20, 30, 40, 50, 60, 70, 80, 90, and 100 Hz, each delivered in 2-s trains with a 1-s inter-train interval. Findings from these mechanistic sweeps were subsequently applied to volitional tasks: frequencies that preferentially recruited flexors or extensors at rest were used during attempted wrist flexion and extension to test whether frequency-dependent biases could enhance task-relevant motor output.

### Functional tasks

Participants performed wrist flexion-extension, grip, and tripod pinch to assess the functional relevance of cervical ESS. For wrist flexion-extension, the forearm was secured in a neutral position using padded restraints mounted on a custom-built rigid frame designed to minimize compensatory movements of the shoulder and trunk. Grip strength was measured with a custom dynamometer incorporating an LSB200 load cell (Futek, USA) embedded in a 3D-printed housing and connected to an amplifier interface, allowing isometric hand closure while restricting wrist deviation. Tripod pinch was assessed with a custom device featuring three contact surfaces (thumb, index, and middle fingers) within a single casing, coupled to an LSM300 miniature load cell (Futek, USA) for high-resolution force measurement (**Fig. 3e**).

Force signals were sampled at 2,000 Hz and low-pass filtered at 10 Hz. Maximum force per trial was defined as the peak within a 2-s task window, and values from two to three trials were averaged per condition. Concurrent EMG recordings from task-relevant flexor and extensor muscles confirmed selective recruitment and agonist-antagonist modulation during stimulation. Comparisons were made across three conditions: No ESS and Optimized ESS, enabling evaluation of whether stimulation enhanced task-specific voluntary performance.

To determine torque production over the stimulation conditions, the raw force vectors (**F** = [F_x_,F_y_,F_z_]) were downsampled to 100 Hz. To account for static loading and sensor bias, a baseline drift correction was applied to each stimulation trial by subtracting the mean force recorded during the pre-stimulation period from the subsequent samples such that

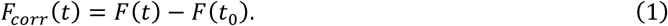

Corrected force vectors were transformed into sensor-frame torque (τ_s_) via the cross product of the known moment arm (r) and the force vector

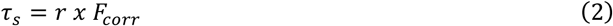

where r = [5.34, −23.99, −6.38] mm. To resolve these torques into the relevant anatomical frame, flexion/extension (FE), supination/pronation (SP), and ulnar/radial deviation (URD), the sensor-frame torque was projected using the transformation matrix (T_a_) by

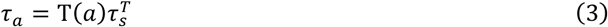

where T_a_ was empirically derived by applying known forces in specific anatomical directions to the measurement device and measuring the resultant output.

To optimize the separation of the anatomical frame and to correct for any sensor misalignment relative to the true physiological axes, we employed an orthogonal rotation based on principle component analysis (PCA). All steady-state torque vectors across the stimulation trials were aggregated into a global variance matrix. PCA was then used to identify the eigenvectors, or principal components, that maximized the variance within the dataset.

The first and second principal components were assigned to the primary movement axes (FE and SP) used for this analysis. To ensure that the PCA-derived axes remained consistent with the anatomical definitions, each new eigenvector (v_pc_) was validated against the original empirical matrix (T_a,i_) using the dot product where

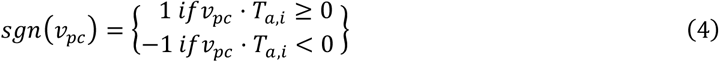

which ensured that the positive direction of the PCA optimized axes continued to represent the intended axes (i.e., FE or SP).

### ESS optimization strategy

To systematically identify stimulation programs that maximized functional output while maintaining selectivity, we applied a structured decision-making framework based on three adjustable domains: location, frequency, and intensity (**Supplementary Figure 1**).

#### Location

Motor pool mapping was used to define segmental preferences of task-relevant muscles. Wrist flexors (FCR, FCU) were most consistently recruited from C7–C8 contacts, whereas wrist extensors (ECR, ECU) were preferentially engaged from C6–C7. For functional enhancement, electrode configurations were systematically biased rostrally (toward C6–C7) when the goal was to facilitate wrist extension, and caudally (toward C7–C8) when stabilizing wrist flexion, grip, or pinch. Small rostrocaudal shifts of just one to two contacts often reversed the flexor–extensor balance, requiring reassessment of EMG responses after each adjustment.

#### Frequency

Once location was optimized, stimulation frequency was adjusted to shape agonist–antagonist recruitment. Low frequencies (≤20–30 Hz) biased flexor dominance and were useful for probing circuitry or stabilizing grip/pinch via co-contraction, but tended to suppress extensors. Mid-range frequencies (30–50 Hz) produced more balanced activation. High frequencies (60–70 Hz) facilitated extensors by promoting temporal summation, thereby overcoming the inhibitory dominance of flexors and enabling robust wrist extension. If high frequency alone did not produce extensor recruitment, intensity was increased (up to 1.5–5.0× MT). Conversely, if low frequency induced excessive flexor tone, intensity was reduced or the electrode configuration shifted rostrally.

#### Intensity

Stimulation amplitude was titrated at rest to determine motor threshold (MT), defined as the lowest current eliciting consistent EMG activity in the target muscle across ≥50% of trials. Intensity was then confirmed during attempted volitional movements. Sub-motor currents (<0.8× MT) produced only sensory priming without measurable facilitation. Intensities around 1.0–1.2× MT reliably engaged propriospinal and interneuronal networks supporting task-specific activation. Higher amplitudes (up to 5.0× MT) broadened recruitment but risked diffuse co-contraction, often biased toward flexor dominance; in such cases, intensity was reduced until task-specific selectivity was restored.

This structured approach allowed electrode location, frequency, and amplitude to be adjusted stepwise in a physiologically guided manner, enabling reproducible targeting of task-relevant motor pools for both mechanistic testing and functional tasks.

### Data processing and statistical analyses

Analyses were performed using MATLAB R2024b (MathWorks) and OriginPro 2025 (OriginLab). Data normality was assessed using the Shapiro-Wilk test. Differences between conditions (No ESS and Optimized ESS) were evaluated using paired two-tailed Student’s t-tests (for normally distributed data) or Wilcoxon signed-rank tests (for non-parametric data). Bonferroni corrections were applied for multiple pairwise comparisons. Statistical significance was defined as P < 0.05. For frequency-dependent modulation and functional force generation, EMG root mean square (RMS) and peak force values were averaged across trials and compared.

## Data Availability

All data supporting the findings of this study are available within the paper and its supplementary information files. Source data for the figures and tables are available from the corresponding author upon reasonable request.

## Data availability

Source data are provided with this paper.

## Code availability

All software and code used to generate the figures in this manuscript are available upon reasonable request to the corresponding author.

## Acknowledgments

This work was supported by philanthropic funding from Paula and Joseph C. “Rusty” Walter III, the Walter Oil & Gas Corporation, and the NeuroSpark seed fund. Additional funding was provided by the National Institutes of Health (NIH) grant 5R01NS119587 and the Craig H. Neilsen Foundation (Research Grant 733278). The funders had no role in the study design, data collection, analysis, or interpretation, nor in the decision to write or submit this article. The authors extend their sincere gratitude to the Houston Methodist Research Institute’s Office of Regulatory Affairs and the operating room personnel for their invaluable technical and clinical assistance during the procedures. We are also grateful to Dr. Gillian Hamilton for her insightful contributions to the writing and proofreading of the manuscript. Most importantly, we deeply appreciate the dedication, motivation, and perseverance of the research participants, without whom this work would not have been possible. We further acknowledge the late Dr. V. Reggie Edgerton for his foundational scientific guidance and pioneering contributions to the field of spinal cord neuromodulation.

## Author contributions

D.G.S. and P.J.H. conceived the study and secured funding. D.G.S., J.O., A.G.S., M.S., and C.M. designed the experiments and assessment protocols. M.M. provided guidance on the stimulation control system. J.D., A.S., and R.K. managed participant recruitment and clinical monitoring. A.H.F. and D.G.S. co-designed the neurosurgical approach, and A.H.F. performed the surgical procedures. J.O., A.G.S., M.S., C.M., V.A.D., and A.V.P. implemented the experiments. J.O., A.G.S., and J.S. analyzed the data and performed statistical evaluations. J.O. and A.G.S. prepared the figures. C.K., T.M.H., and Y.F. served on the Data and Safety Monitoring Board. The manuscript was written by D.G.S., J.O., A.G.S., and M.S., with revisions and final approval from all authors.

## SUPPLEMENTARY FIGURES

**Supplementary Figure 1.**
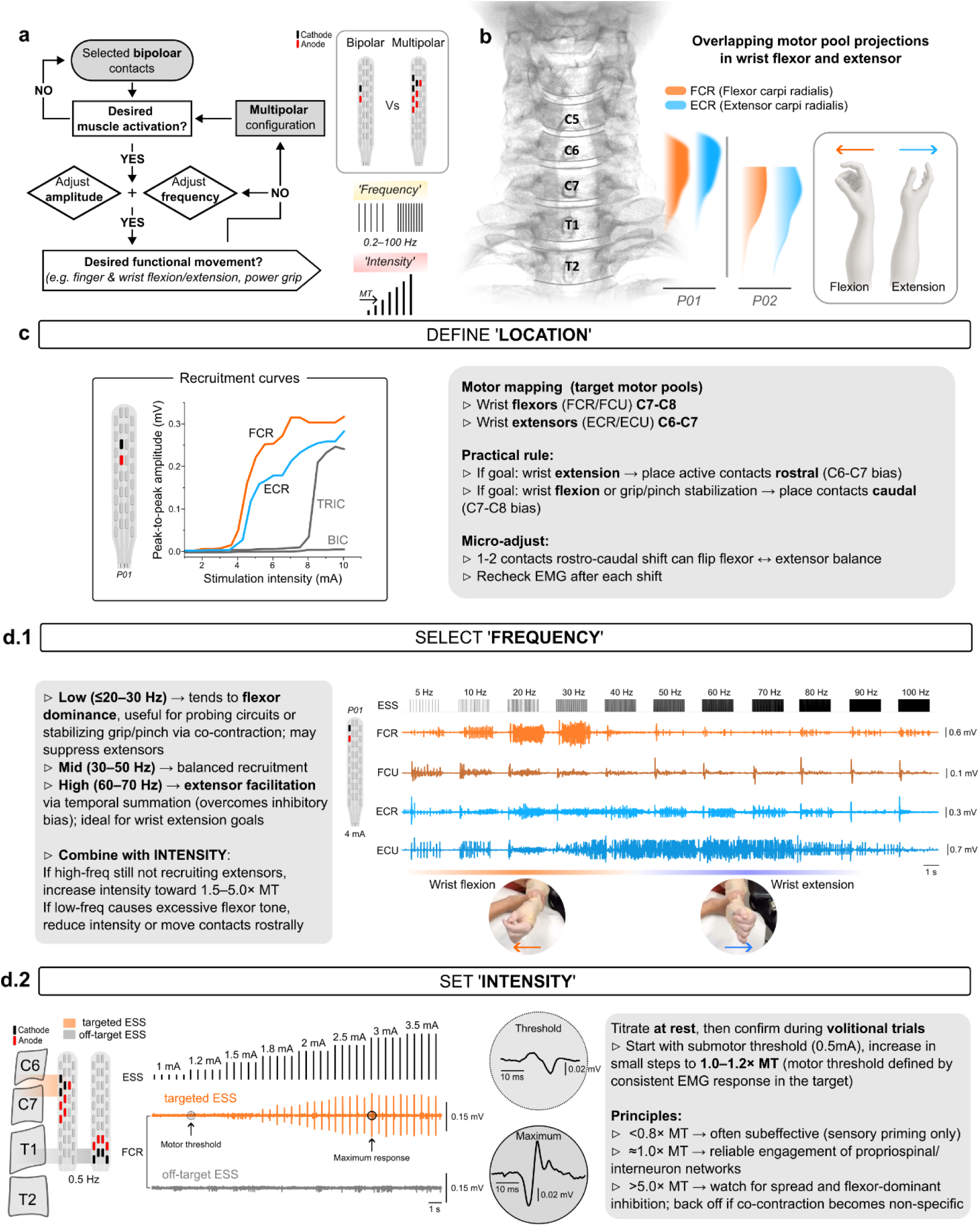
Structured decision framework for ESS optimization. **a.** Flowchart illustrating the iterative decision-making process for optimizing cervical epidural spinal stimulation (ESS). The framework guides the systematic tuning of electrode configuration (bipolar vs. multipolar), frequency, and amplitude to achieve desired functional movements. **b.** Anatomical overlay of overlapping motor pool projections for wrist flexors (FCR, orange) and extensors (ECR, blue) across the cervical spinal cord (C5–T2) in P01 and P02. The dense anatomical overlap necessitates parameter tuning beyond spatial targeting alone. **c.** Define ‘Location’. Representative recruitment curves (P01) identify segmental preferences. A clinical heuristic was established: rostral active contacts (C6–C7) preferentially engaged wrist extensors, whereas caudal contacts (C7–C8) favored wrist flexors. **d.1.** Select ‘Frequency’. Representative electromyography (EMG) traces demonstrate frequency-dependent modulation. Low frequencies (≤20-30 Hz) promoted wrist flexors dominance, mid-range frequencies (30-50 Hz) yielded balanced recruitment, and high frequencies (60-70 Hz) facilitated activation of wrist extensors. **d.2.** Set ‘Intensity’. Representative EMG traces demonstrate amplitude titration from sub-motor threshold (MT) to suprathreshold levels. Functional engagement was typically initiated near 1.0-1.2×MT, while higher intensities (>5.0×MT) were often required to maximize force production and reliably recruit extensors. These optimized parameters maximized selective motor output while limiting the spread of activation that leads to non-specific co-contraction. Note that ESS targeted to specific motor pools elicited graded recruitment with clear thresholds and saturation at higher intensities, whereas off-target ESS, delivered through electrode contacts outside functionally relevant segments, produced minimal or inconsistent activation, serving as a control to dissociate non-specific stimulation effects.

**Supplementary Figure 2.**
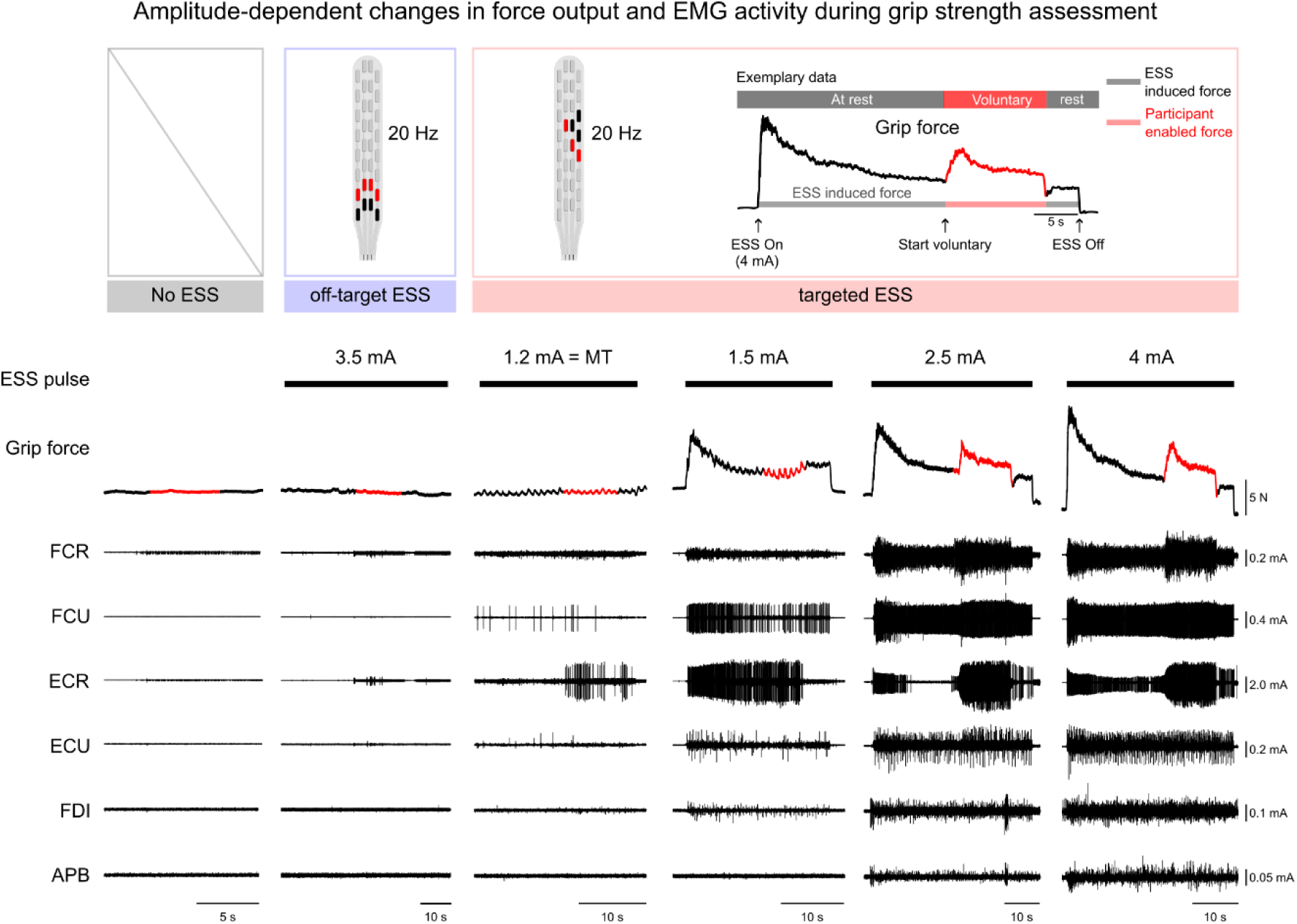
Location- and amplitude-dependent modulation of grip strength. Top: Experimental paradigm and conditions overview. Representative data illustrates the assessment protocol: stimulation delivered at a segment-specific location and intensity above motor threshold (MT) induces and increase in resting force (black trace; ESS-induced force), followed by a voluntary increase in grip force reflecting participant-generated output (red trace). Bottom: Raw grip force and electromyography (EMG) traces across varying stimulation locations and intensities. Off-target ESS, delivered through electrode contacts outside functionally relevant segments, produced minimal activation even at higher intensities (3.5 mA). By contrast, during segment-specific (targeted) ESS, stimulation intensities substantially above MT were required to achieve coordinated recruitment of forearm and intrinsic hand muscles, with functionally robust force generation emerging only at suprathreshold intensities (2.5–4.0 mA). These findings indicate that functional enhancement depends on the integration of spatial targeting and amplitude titration.

**Supplementary Figure 3.**
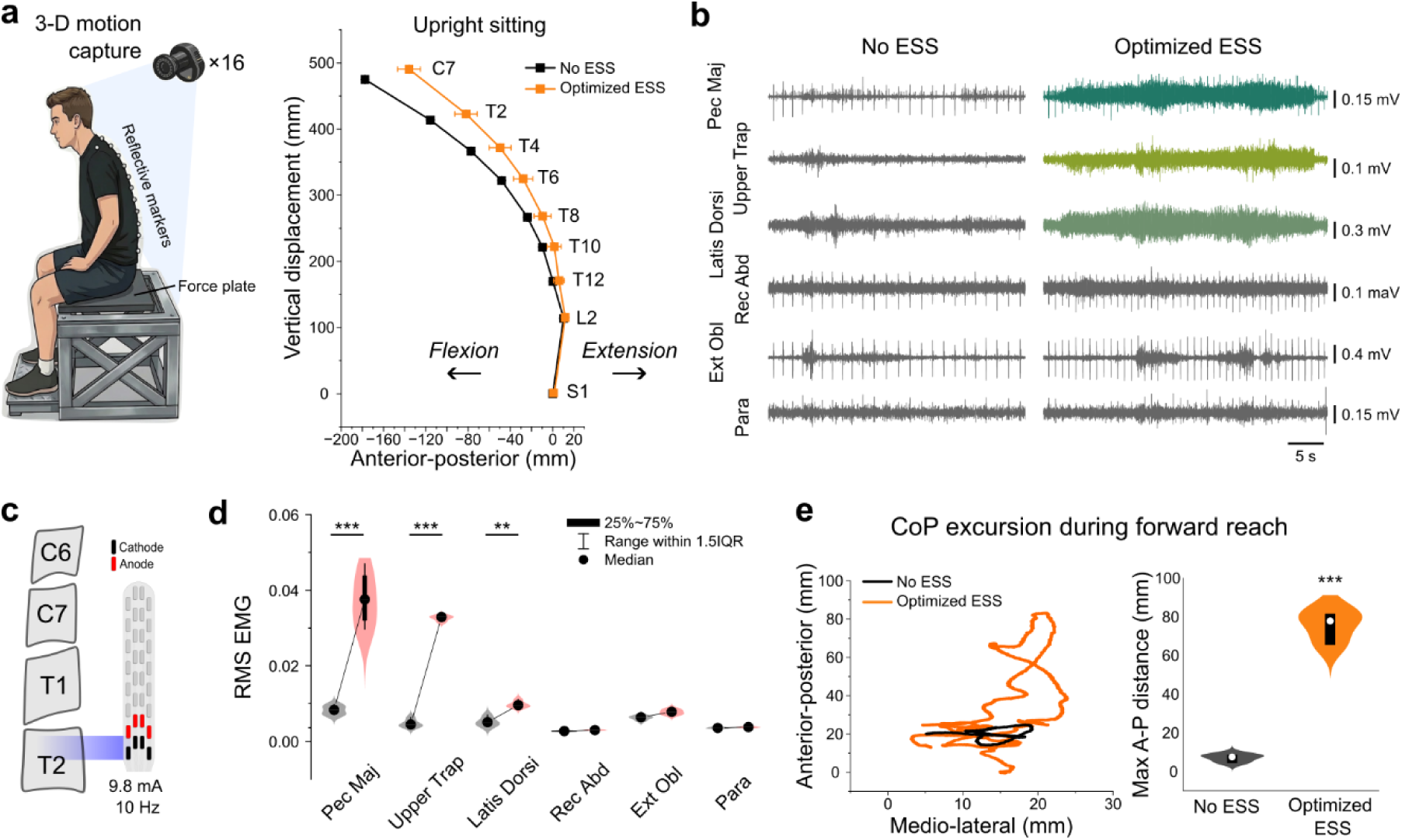
Immediate improvement in trunk stability and seated balance control during ESS. **a.** Kinematic assessment of upright sitting posture in P02. Left: Schematic of the 3D motion capture and force plate setup. Right: Spatial coordinates of spinal markers (C7 to S1) in the sagittal plane. Optimized ESS (orange) reduced abnormal anterior flexion compared to the No ESS condition (black), restoring a more erect and stable trunk posture. **b.** Representative electromyography (EMG) traces of trunk musculature during unsupported sitting. Optimized ESS elicited robust, sustained activation of upper trunk and proximal muscles (pectoralis major, upper trapezius, and latissimus dorsi) compared to the No ESS condition. **c.** Optimized ESS utilized caudal contacts at the T2 level, specifically selected to facilitate trunk stability and postural control (9.8 mA, 10 Hz). **d.** Quantitative analysis of root mean square (RMS) EMG activity. Violin plots highlight stimulation-driven increases in upper trunk muscle activation. **e.** Center of Pressure (CoP) excursion during a dynamic forward reaching task. Left: Representative CoP trajectories. Right: Violin plots quantifying maximum anterior-posterior (A-P) CoP distance. Optimized ESS (orange) enabled significantly greater dynamic CoP excursion compared to No ESS (black), reflecting enhanced postural control and balance during functional reach. Box plots (d, e) within the violins display the median (circle), interquartile range (thick bar, 25th–75th percentiles), and whiskers extending to 1.5× the interquartile range. Asterisks denote statistical significance (**P < 0.01, ***P < 0.001; see Methods for details of statistical analyses). Pec Maj: pectoralis major, Upper Trap: upper trapezius, Latis Dorsi: latissimus dorsi, Rec Abd: rectus abdominis, Ext Obl: external oblique, Para: paraspinals.

**Supplementary Figure 4.**
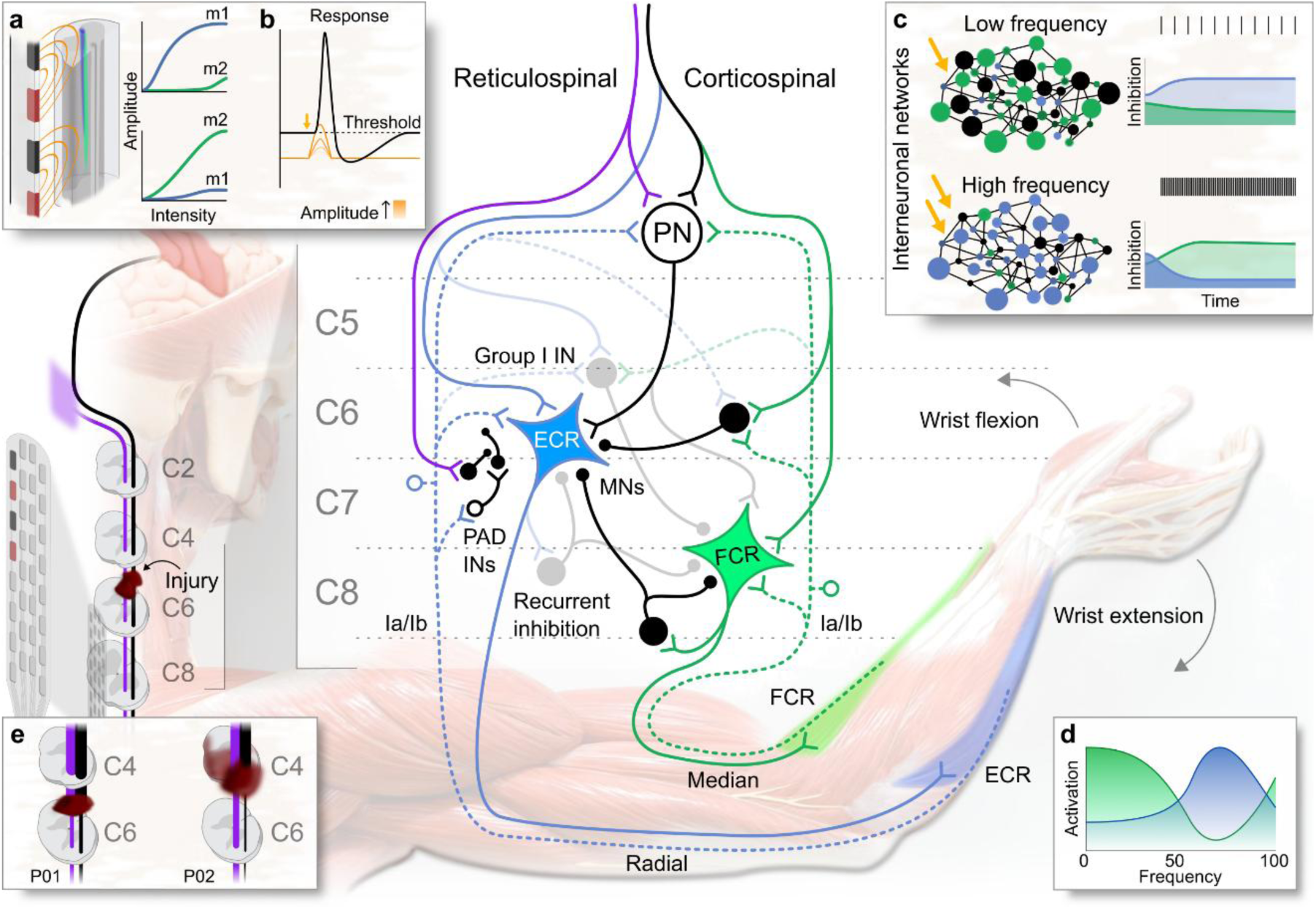
Presumed spinal and supraspinal pathways mediating parameter-specific cervical ESS. The schematic illustrates segmental interneuronal circuits and descending pathways plausibly recruited by targeted ESS to engage overlapping flexor carpi radialis (FCR, green) and extensor carpi radialis (ECR, blue) motoneuron pools. **a, b**. Schematic of spatial location- (**a**) and intensity-dependent (**b**) recruitment of distinct motor pools. **c, d**. Proposed mechanisms of frequency-dependent modulation. Low-frequency stimulation (top, **c**) preferentially engages FCR-associated Ia afferents and inhibitory interneurons, biasing the network toward flexor dominance (green shading, **d**). Conversely, high-frequency stimulation (bottom, **c**) facilitates temporal summation within premotoneuronal circuits, overcoming flexor-biased inhibition to elicit disynaptic excitation of ECR motoneurons and promote wrist extension (blue shading, **d**). During attempted voluntary movement, residual corticospinal drive converges with ESS-evoked afferent input on propriospinal neurons (PN) and local interneurons (e.g., PAD INs, Group I INs, and those mediating recurrent inhibition). **E**. Influence of lesion anatomy. Following cervical spinal cord injury (SCI), the loss of descending control over presynaptic inhibition, combined with an altered balance of reticulospinal (purple) versus corticospinal (black) influence, may bias segmental processing toward flexor synergies. This flexor dominance is predicted to be pronounced in injuries with more rostral or extensive lesions (e.g., P02 vs. P01).

## References

1. Center, N.S.C.I.S. “Traumatic spinal cord injury facts and figures at a glance”. Birmingham, AL: University of Alabama at Birmingham. Available online. (2024).

2. Anderson, K.D. Targeting recovery: priorities of the spinal cord-injured population. J. Neurotrauma 21, 1371–1383 (2004).

3. Thorogood, N.P., et al. Spinal Cord Injury Community Personal Opinions and Perspectives on Spinal Cord Stimulation. Top. Spinal Cord Inj. Rehabil. 29, 1–11 (2023).

4. Taccola, G., Sayenko, D., Gad, P., Gerasimenko, Y. & Edgerton, V.R. And yet it moves: Recovery of volitional control after spinal cord injury. Prog. Neurobiol. 160, 64–81 (2018).

5. Chalif, J.I., et al. Epidural Spinal Cord Stimulation for Spinal Cord Injury in Humans: A Systematic Review. J. Clin. Med. 13 (2024).

6. Taylor, C., et al. Transcutaneous spinal cord stimulation and motor responses in individuals with spinal cord injury: A methodological review. PLoS One 16, e0260166 (2021).

7. Scheffler, M.S., Martin, C.A., Dietz, V., Faraji, A.H. & Sayenko, D.G. Synergistic implications of combinatorial rehabilitation approaches using spinal stimulation on therapeutic outcomes in spinal cord injury. Clin. Neurophysiol. 165, 166–179 (2024).

8. Gill, M.L., et al. Neuromodulation of lumbosacral spinal networks enables independent stepping after complete paraplegia. Nat. Med. 24, 1677–1682 (2018).

9. Rowald, A., et al. Activity-dependent spinal cord neuromodulation rapidly restores trunk and leg motor functions after complete paralysis. Nat. Med. 28, 260–271 (2022).

10. Angeli, C.A., et al. Recovery of over-ground walking after chronic motor complete spinal cord injury. N. Engl. J. Med. 379, 1244–1250 (2018).

11. Greiner, N., et al. Recruitment of upper-limb motoneurons with epidural electrical stimulation of the cervical spinal cord. Nat. Commun. 12, 435 (2021).

12. Inanici, F., Brighton, L.N., Samejima, S., Hofstetter, C.P. & Moritz, C.T. Transcutaneous spinal cord stimulation restores hand and arm function after spinal cord injury. IEEE Trans. Neural Syst. Rehabil. Eng. **PP**(2021).

13. Moritz, C., et al. Non-invasive spinal cord electrical stimulation for arm and hand function in chronic tetraplegia: a safety and efficacy trial. Nat. Med. 1–8 (2024).

14. Gad, P., et al. Non-Invasive Activation of Cervical Spinal Networks after Severe Paralysis. J. Neurotrauma 35, 2145–2158 (2018).

15. de Freitas, R.M., et al. Selectivity and excitability of upper-limb muscle activation during cervical transcutaneous spinal cord stimulation in humans. J. Appl. Physiol. (1985) 131, 746–759 (2021).

16. Oh, J., et al. Cervical transcutaneous spinal stimulation for spinal motor mapping. iScience 25, 105037 (2022).

17. Lu, D.C., et al. Engaging Cervical Spinal Cord Networks to Reenable Volitional Control of Hand Function in Tetraplegic Patients. Neurorehabil. Neural Repair 30, 951–962 (2016).

18. Freyvert, Y., et al. Engaging cervical spinal circuitry with non-invasive spinal stimulation and buspirone to restore hand function in chronic motor complete patients. Sci. Rep. 8, 15546 (2018).

19. Powell, M.P., et al. Epidural stimulation of the cervical spinal cord for post-stroke upper-limb paresis. Nat. Med. 29, 689–699 (2023).

20. Fawcett, J. W., et al. Guidelines for the conduct of clinical trials for spinal cord injury as developed by the ICCP panel: spontaneous recovery after spinal cord injury and statistical power needed for therapeutic clinical trials. Spinal Cord, 45(3), 190–205 (2007).

21. Jenny, A. & Inukai. Principles of motor organization of the monkey cervical spinal cord. J. Neurosci. 3, 567–575 (1983).

22. Milosevic, M., Masugi, Y., Sasaki, A., Sayenko, D.G. & Nakazawa, K. On the reflex mechanisms of cervical transcutaneous spinal cord stimulation in human subjects. J. Neurophysiol. 121, 1672–1679 (2019).

23. Mahan, E.E., et al. Assessing the Effect of Cervical Transcutaneous Spinal Stimulation With an Upper Limb Robotic Exoskeleton and Surface Electromyography. IEEE Trans. Neural Syst. Rehabil. Eng. 32, 2883–2892 (2024).

24. Barra, B., et al. Epidural electrical stimulation of the cervical dorsal roots restores voluntary upper limb control in paralyzed monkeys. Nat. Neurosci. 25, 924–934 (2022).

25. Tuazon, J.R., Jahan, A., Jutai, J.W. Understanding adherence to assistive devices among older adults: a conceptual review. Disabil. Rehabil. Assist. Technol. (2019).

26. Almeida, S.B., et al. Assistive technology in spinal cord injury rehabilitation: use or non-use? Understanding what happens post-discharge in tetraplegic individuals at a rehabilitation centre in Northeast Brazil. Disabil. Rehabil. Assist. Technol. 20, 1700–1710 (2025).

27. Routledge, N., Zhang, D. & Metcalfe, B. Electrical stimulation in upper limb assistance: opportunities and challenges. Front. Neurosci. 19, 1702889 (2025).

28. Hultborn, H., Meunier, S., Pierrot-Deseilligny, E. & Shindo, M. Changes in presynaptic inhibition of Ia fibres at the onset of voluntary contraction in man. J. Physiol. 389, 757–772 (1987).

29. Pierrot-Deseilligny, E. & Burke, D. The Circuitry of the Human Spinal Cord: Its Role in Motor Control and Movement Disorders (Cambridge Univ. Press, 2005).

30. Kendall, F.P. Muscles : testing and function with posture and pain, (Lippincott Williams & Wilkins, Baltimore, MD, 2005).

31. Burke, D., Gandevia, S.C. & McKeon, B. The afferent volleys responsible for spinal proprioceptive reflexes in man. J. Physiol. 339, 535–552 (1983).

32. Aimonetti, J.-M., Vedel, J.-P., Schmied, A. & Pagni, S. Inhibition versus facilitation of the reflex responsiveness of identified wrist extensor motor units by antagonist flexor afferent inputs in humans. Exp. Brain Res. 133, 391–401 (2000).

33. Taccola, G., et al. Interactions between descending and spinal circuits on motor output. Exp. Neurol. 392, 115347 (2025).

34. McIntosh, J.R., et al. Timing-dependent synergies between motor cortex and posterior spinal stimulation in humans. J. Physiol. 602, 2961–2983 (2024).

35. Mizuno, Y., Tanaka, R. & Yanagisawa, N. Reciprocal group I inhibition on triceps surae motoneurons in man. J. Neurophysiol. 34, 1010–1017 (1971).

36. Alstermark, B. & Isa, T. Circuits for skilled reaching and grasping. Annu. Rev. Neurosci. 35, 559–578 (2012).

37. Alstermark, B., Isa, T., Pettersson, L.G. & Sasaki, S. The C3-C4 propriospinal system in the cat and monkey: a spinal pre-motoneuronal centre for voluntary motor control. Acta Physiol. (Oxf.) 189, 123–140 (2007).

38. Sangari, S. & Perez, M.A. Distinct Corticospinal and Reticulospinal Contributions to Voluntary Control of Elbow Flexor and Extensor Muscles in Humans with Tetraplegia. J. Neurosci. 40, 8831–8841 (2020).

39. Baker, S.N. & Perez, M.A. Reticulospinal Contributions to Gross Hand Function after Human Spinal Cord Injury. J. Neurosci. 37, 9778–9784 (2017).

40. Bunday, K.L. & Perez, M.A. Motor recovery after spinal cord injury enhanced by strengthening corticospinal synaptic transmission. Curr. Biol. 22, 2355–2361 (2012).

41. American Spinal Injury Association. International Standards for the Neurological Classification of Spinal Cord Injury (ISNCSCI). (2019).

42. Sayenko, D.G., et al. Spinal segment-specific transcutaneous stimulation differentially shapes activation pattern among motor pools in humans. J. Appl. Physiol. 118, 1364–1374 (2015).

43. Calvert, J.S., et al. Electrophysiological Guidance of Epidural Electrode Array Implantation over the Human Lumbosacral Spinal Cord to Enable Motor Function after Chronic Paralysis. J. Neurotrauma 36, 1451–1460 (2019).

